# Behavioural barriers to COVID-19 testing in Australia: Two national surveys to identify barriers and estimate prevalence by health literacy level

**DOI:** 10.1101/2021.08.26.21262649

**Authors:** Carissa Bonner, Carys Batcup, Julie Ayre, Kristen Pickles, Erin Cvejic, Tessa Copp, Samuel Cornell, Rachael Dodd, Jennifer Isautier, Brooke Nickel, Kirsten McCaffery

**Affiliations:** Sydney Health Literacy Lab, School of Public Health, Faculty of Medicine and Health, The University of Sydney

## Abstract

**Background:** COVID-19 testing and contact tracing has been crucial in Australia’s prevention strategy. However, testing for COVID-19 is far from optimal, and behavioural barriers are unknown. Study 1 aimed to identify the range of barriers to testing. Study 2 aimed to estimate prevalence in a nationally relevant sample to target interventions.

**Methods:** Study 1: National longitudinal COVID-19 survey from April-November 2020. Testing barriers were included in the June survey (n=1369). Open responses were coded using the COM-B framework (capability-opportunity-motivation). Study 2: Barriers from Study 1 were presented to a new nationally representative sample in November to estimate prevalence (n=2869). Barrier prevalence was analysed by health literacy level using Chi square tests.

**Results:** Study 1: 49% strongly agreed to get tested for symptoms, and 69% would self-isolate. Concern about pain was the top barrier from a provided list (11%), but 32 additional barriers were identified from open responses and coded to the COM-B framework. Study 2: The most prevalent barriers were motivation issues (e.g. don’t believe symptoms are COVID-19: 28%, few local cases: 18%). Capability issues were also common (e.g. not sure symptoms are bad enough: 19%, not sure whether symptoms need testing: 15%). Many barriers were more prevalent amongst people with low compared to high health literacy, including motivation (preference to self isolate: 21% vs 12%, pain: 15% vs 9%) and capability (not sure symptom needs testing: 12% vs 8%, not sure how to test:11% vs 4%).

**Conclusion:** Even in a health system with free and widespread access to COVID-19 testing, motivation and capability barriers were prevalent issues, particularly for people with lower health literacy. This study highlights the important of diagnosing behaviour barriers to target public health interventions for COVID-19 and future pandemics.

## BACKGROUND

Individual behaviour is crucial to the control of COVID-19, but research on non-pharmaceutical strategies for COVID-19 prevention has been underfunded^1^. The first step to develop such strategies is diagnosing what behavioural barriers must be addressed to increase compliance with COVID-19 prevention. A key prevention behaviour that has been overlooked in proposed models of COVID-19 prevention is COVID-19 testing^2^.

The successful COVID-19 prevention strategy in Australia has been dependent on people getting tested when they have been in contact with a case, or have COVID-19 symptoms (e.g. fever, cough, sore throat). Community members must then self-isolate at home until they return a negative result. This test-trace-isolate strategy has been used to determine the need for short-term localised restrictions until clusters are brought under control^3^. According to the COM-B model^4^, such COVID-19 prevention behaviours can be conceptualised in terms of three main drivers: physical/psychological *capability* (e.g. having the physical ability to drive to and walk up the stairs to access a testing centre, and knowing what to do if you have symptoms), physical/social *opportunity* (e.g. the availability of testing centres in your area, and social norms that make self-isolation acceptable), and automatic/reflective *motivation* (e.g. getting into the habit of staying home when you have symptoms, and believing that it’s important to get tested if you develop symptoms)^2^.

There is little research on COVID-19 testing behaviours given the very new nature of this issue, but emerging literature and media reports suggest different barriers exist across countries. For example, countries such as Tanzania have major issues with opportunity barriers in terms of limited access to COVID-19 tests and fake testing kits^5^. Cost may be a barrier in other countries such as the UK and US where health insurance may not cover the testing. This disproportionately affects certain groups such as immigrant and non-citizen communities, who may also fear financial and legal repercussions from testing positive^6^. Testing may be limited to certain criteria (e.g. only if you have symptoms, regardless of exposure to COVID-19 cases) due to lack of supply or staff resource issues^7^. There may also be issues of delivering tests and transporting samples for remote areas^8^. Finally, there may be issues with inadequate communication and low community knowledge about which symptoms require testing and the process to follow^7^.

In terms of opportunity barriers, Australia is fortunate to have efficient and free testing widely available, although this can vary by location. Testing clinics have been set up all around the country, which includes drive through options to minimise contact with others, and results are generally sent by text message within 2 days^9,10^. However, the process can take longer and is unpredictable where demand increases in new outbreak areas such as Victoria in July-September 2020^11–13^.

For capability barriers, public health messages have focused on basic processes but we know certain groups are less likely to understand and act on COVID-19 prevention advice. For example in Australia, people with lower health literacy and those who speak a language other than English were less likely to be able to identify COVID-19 symptoms and prevention measures^14^. Younger people, men and those with less education were more likely to agree with COVID-19 misinformation about prevention, management and cure^15^. These groups likely need different communication strategies to ensure everyone *understands* the message and takes appropriate *action*.

To address motivation, messaging trials have explored different strategies to increase intentions to follow COVID-19 prevention advice, e.g. framing the behaviour as helping the community rather than avoiding individual risk^16^, or testing identity-based messages such as “don’t be a spreader”^17^. However, emerging research in this area has neglected testing as a key behavioural outcome for COVID-19 prevention. We need to understand the behavioural barriers^2^ to testing in order to develop and evaluate interventions to address these issues for managing COVID-19 more effectively now. With new variants and unequal access, the vaccine rollout is not guaranteed to eliminate COVID-19 in the near future, so we will be reliant on testing for many months and even years to come^18^. Better understanding of testing barriers is a pressing issue for COVID-19, but also essential to improve the management of future pandemics.

## AIM

We conducted two Australian studies to address a major gap in our understanding about how to improve COVID-19 prevention. Study 1 aimed to identify the *range* of barriers to COVID-19 testing. Study 2 aimed to estimate the *prevalence* of barriers to COVID-19 testing, to target interventions to the most important issues.

## METHOD

### Study 1: Barrier identification

The Sydney Health Literacy Lab (SHeLL) has been conducting a national longitudinal survey in Australia since April 2020^14,19^. The original sample was recruited via an online market research panel, supplemented with social media advertising (n=4326). The social media users were followed-up monthly from April-July. A list of possible testing barriers provided to the authors by the NSW Department of Health was included in the June 2020 survey (n=1369), along with intentions to test and self-isolate if symptomatic. These were worded as: “over the next 4 weeks, I plan to get tested if I have COVID-19 symptoms (cough, sore throat, fever)” and “over the next 4 weeks, I plan to stay home if I have COVID-19 symptoms (cough, sore throat, fever)”; with 1-7 response options from strongly disagree to strongly agree. Open responses to “other” test barriers were also collected. Descriptive analyses are reported as percentages for the survey results. The test barriers identified from the survey responses were categorised using the COM-B framework^4^ by two researchers who have been trained in the use of this framework (CB and CAB). CAB conducted the initial coding, and this was reviewed by CB with discussion of issues that could be coded in different ways until consensus was reached. All barriers could be categorised within the COM-B components.

### Study 2: Barrier prevalence

All barriers identified in Study 1 were presented to a new nationally representative sample in November 2020, recruited through online market research company Dynata with quotas for age, gender, education and state (n=2034). Participants were asked ‘Below are some of the reasons why people don’t want to get tested, or can’t get tested, for COVID-19. Please select the reasons that apply to you. Pick as many or as few as you want. If I get COVID-19 symptoms (signs of having COVID-19), I might not get tested because…’. They were also presented with the option ‘None of these apply to me’. Participants were then prompted to rank their selected barriers in order of importance (‘Now please re-order the answers you gave to the last question, from the most important reason at the top to the least important. Don’t worry about it being exactly right, this will just help us understand broadly which reasons are more important to people.’). We included all barriers identified in study 1 except physical opportunity barriers, as these could not be addressed through a messaging intervention which was the intended use of the results. Comparisons of barrier prevalence between low and high health literacy groups were based on independent Chi square tests. Health literacy was measured using Chew et al.’s validated Health Literacy Screener^20^.

## RESULTS

The sample demographics are shown in Table 1. Study 1 had under-representation of male gender (32%) and lower education (14.5%), with a relatively small sample for low health literacy (8.9%). Quota sampling in Study 2 resulted in a more representative sample for male gender (49.5% male) and lower education (34.6%), which increased the sample for low health literacy (16.5%).

**Table 1:** Descriptive characteristics of samples in Study 1 and Study 2.

| Variable | Study 1<br>(N=1369) | Study 2<br>(n=2034) |
| --- | --- | --- |
| **Age (years), mean (SD)** | 44.7 (16.7) | 45.6 (16.8) |
| **Age group, n (%)** |  |  |
| 18 to 25 years | 232 (16.9%) | 282 (13.9%) |
| 26 to 40 years | 372 (27.2%) | 625 (30.7%) |
| 41 to 55 years | 344 (25.1%) | 502 (24.7%) |
| 56 to 90 years | 421 (30.8%) | 625 (30.7%) |
| **Gender, n (%)** |  |  |
| Male | 433 (31.6%) | 1006 (49.5%) |
| Female | 911 (66.5%) | 1024 (50.3%) |
| Other / prefer not to say | 25 (1.8%) | 4 (0.2%) |
| **Highest level of educational attainment, n (%)** |  |  |
| High school or less | 198 (14.5%) | 704 (34.6%) |
| Certificate I-IV | 140 (10.2%) | 733 (36.0%) |
| University education | 1031 (75.3%) | 597 (29.4%) |
| **Health literacy, n (%)** |  |  |
| Adequate (high) | 1170 (91.1%) | 1699 (83.5%) |
| Inadequate (low) | 199 (8.9%) | 335 (16.5%) |
| **State, n (%)** |  |  |
| Australian Capital Territory | 48 (3.5%) | 41 (2.0%) |
| New South Wales | 721 (52.7%) | 642 (31.6%) |
| Northern Territory | 5 (0.4%) | 19 (0.9%) |
| Queensland | 183 (13.4%) | 422 (20.7%) |
| South Australia | 63 (4.6%) | 140 (6.9%) |
| Tasmania | 55 (4.0%) | 46 (2.3%) |
| Victoria | 205 (15.0%) | 511 (25.1%) |
| Western Australia | 90 (6.6%) | 213 (10.5%) |
| **Region, n (%)** |  |  |
| Rural | 342 (25.0%) | 591 (29.1%) |
| Metropolitan | 1027 (75.0%) | 1443 (70.9%) |

### Study 1: Barrier identification (June 2020, n=1369)

The results of Study 1 survey questions are reported in Table 2. Around half (49.1%) of people strongly agreed they would get tested if they had COVID-19 symptoms (cough, fever, sore throat), but most people agreed to some extent that they would get tested if they had symptoms (83.9%). For self-isolation, 69% strongly agreed they would stay home if they had symptoms. The most common barriers selected from the list provided were that testing is painful (11.2%), not knowing how to get tested (7.1%), and worry about getting infected at the testing centre (5.9%). Many participants (9.9%) indicated other reasons, with 136 open responses that included many additional barriers to testing than those provided in the survey question. Table 3 maps all the barriers identified in open survey responses to the COM-B drivers of behaviour, covering all components.

**Table 2:** Study 1 testing/self-isolation intentions and testing barriers (June 2020, n=1369)

| **Intention to get tested if symptomatic in next 4 weeks** | **n (%)** |
| --- | --- |
| Strongly agree | 674 (49.1%) |
| Agree | 376 (27.4%) |
| Somewhat agree | 101 (7.4%) |
| Neither agree nor disagree | 72 (5.2%) |
| Somewhat disagree | 40 (2.9%) |
| Disagree | 49 (3.6%) |
| Strongly disagree | 60 (4.4%) |
| **Intention to self-isolate if symptomatic in next 4 weeks** | **n (%)** |
| Strongly agree | 951 (69.3%) |
| Agree | 310 (22.6%) |
| Somewhat agree | 53 (3.9%) |
| Neither agree nor disagree | 12 (0.9%) |
| Somewhat disagree | 14 (1.0%) |
| Disagree | 6 (0.4%) |
| Strongly disagree | 26 (1.9%) |
| **Testing barriers (note: multiple options could be selected)** | **n (%)** |
| Testing is painful | 153 (11.2%) |
| Other (please detail) – see Table 3 | 136 (9.9%) |
| I don't know how, when and where to get tested | 98 (7.1%) |
| I'm worried I will get infected with COVID-19 at the testing clinic | 81 (5.9%) |
| I'll forget to get tested | 33 (2.4%) |
| I'm worried about what others think of me | 32 (2.3%) |
| It's too difficult or expensive to get tested | 29 (2.1%) |
| Testing doesn't work / I don't trust the results | 17 (1.2%) |
| No one else is getting tested | 11 (0.8%) |

**Table 3:**
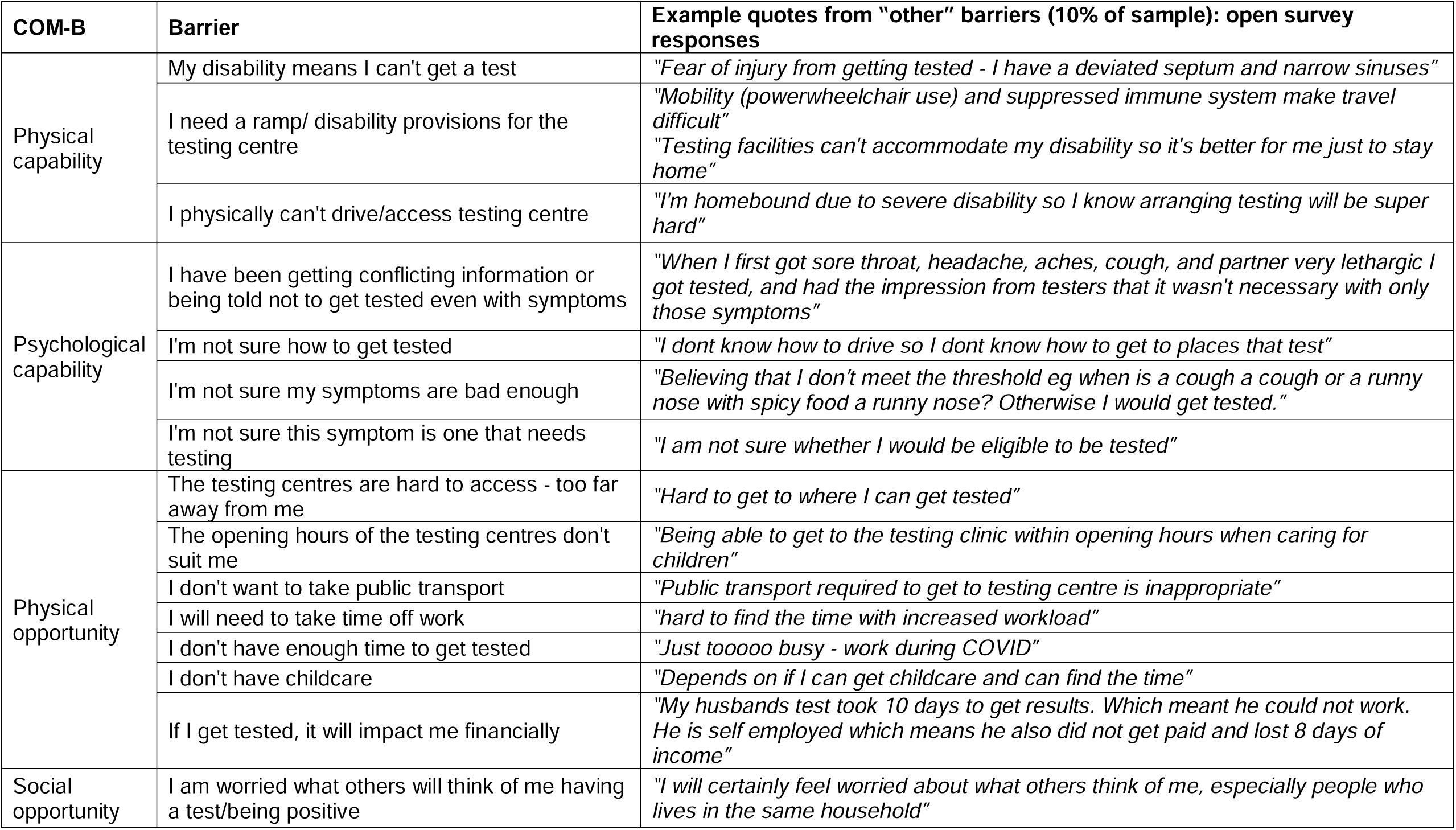
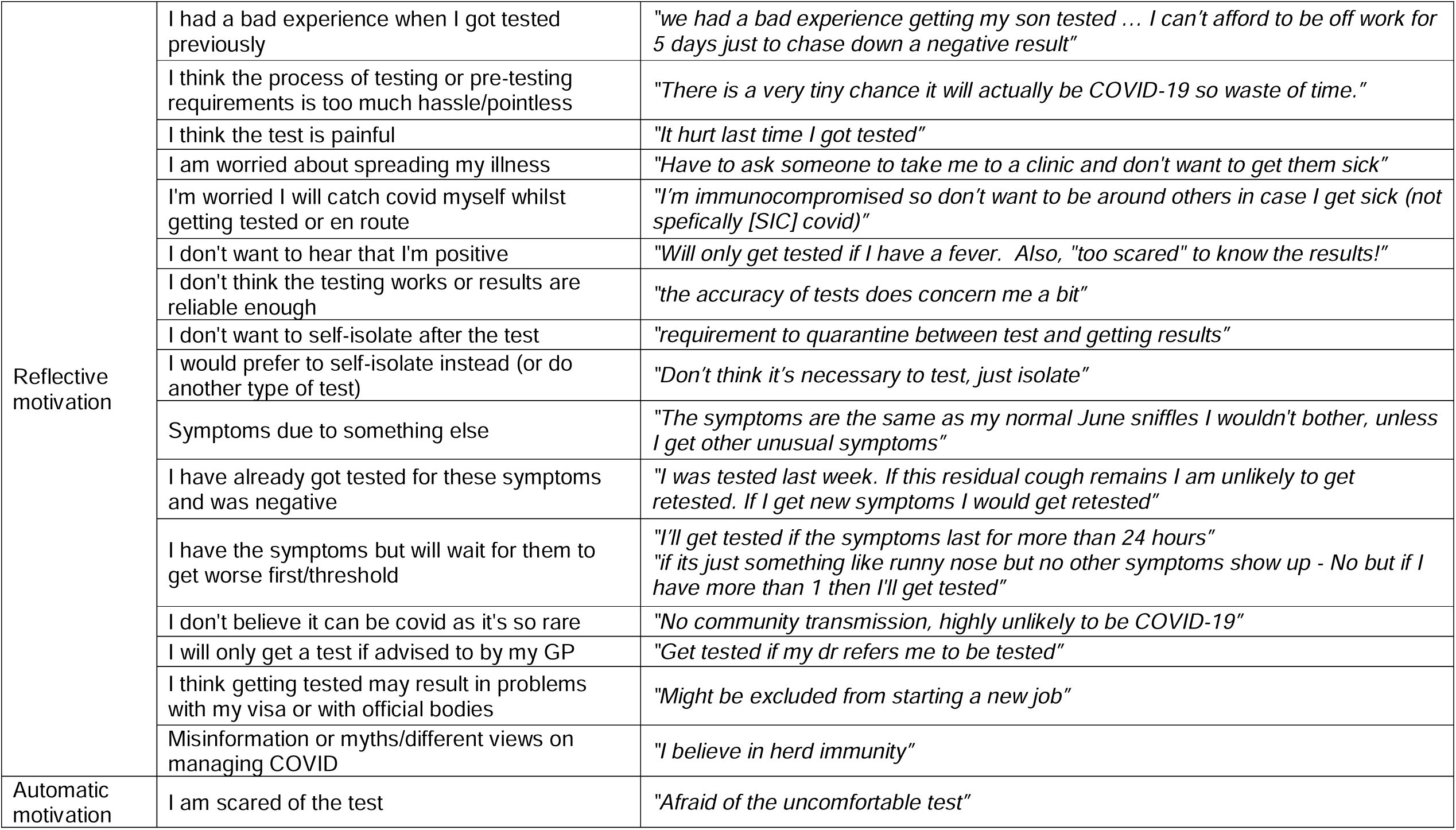
Study 1 COVID-19 testing behaviour barriers mapped to the COM-B model (June 2020)

### Study 2: Barrier prevalence (November 2020, n=2034)

Table 4 shows the prevalence and importance of barriers selected, from those participants who selected any barrier (941, 46.3%). The top barriers were related to motivation: ‘I know what symptoms I have and don’t believe they are COVID-19 ones e.g. hayfever/normal cold’ (selected by 28%, with 10% ranking it most important), and ‘It is unlikely I have COVID-19 because there aren’t many cases in my area’ (18% selected, with 5% ranking it most important). Capability issues were also common: ‘I’m not sure my symptoms are bad enough’ (19% selected, with 5% ranking it most important), and ‘I’m not sure this symptom is one that needs testing” (15% selected, with 3% ranking it most important). Social opportunity issues were uncommon: 6% were worried what others might think if they got a positive COVID-19 test, and 4% worried what others would think if they got tested at all. Physical opportunity issues were not specifically listed as explained in methods, but frequent responses for other issues included disabilities, access (especially distance to travel to a testing centre) and time restrictions.

**Table 4:**
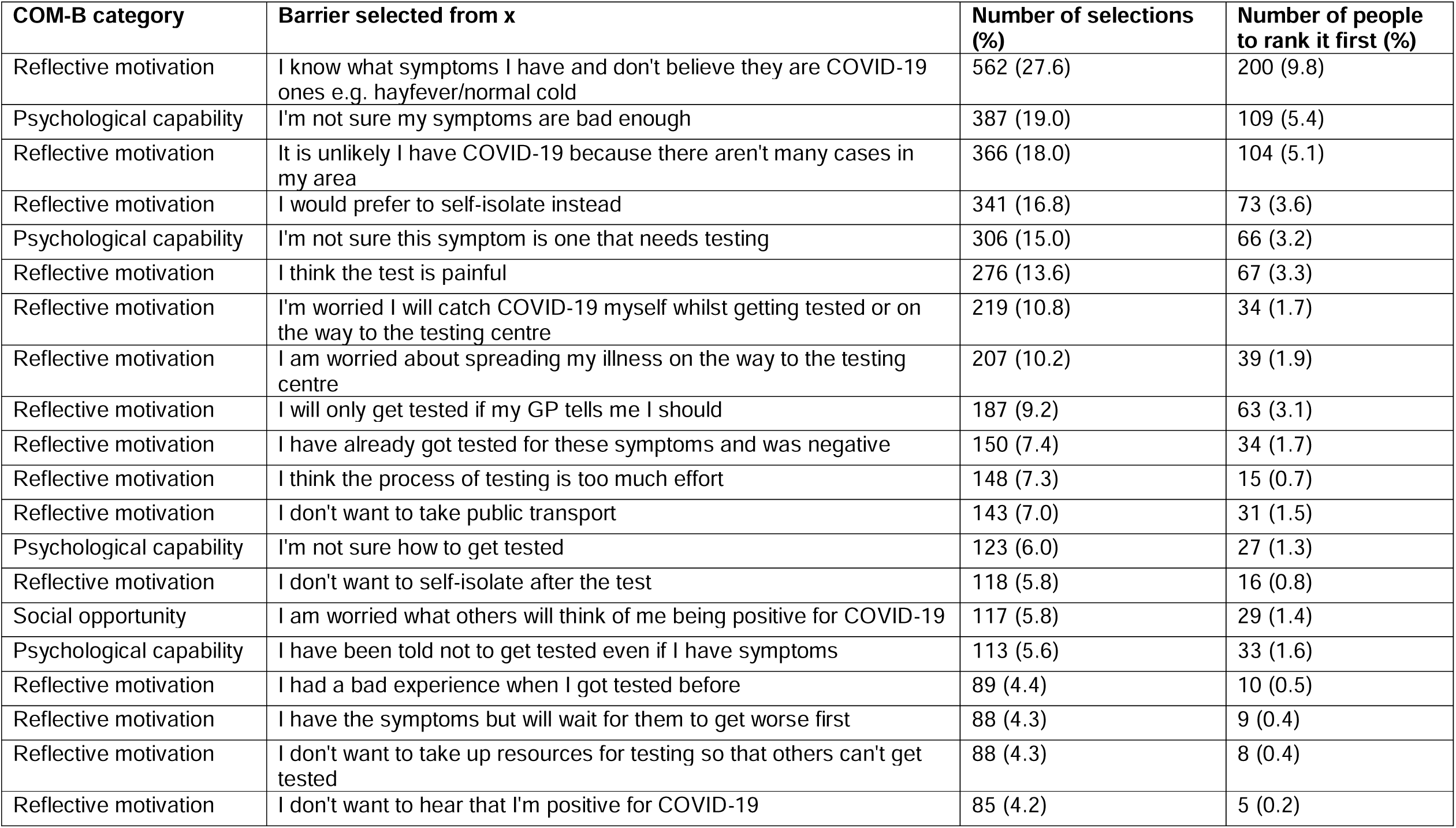
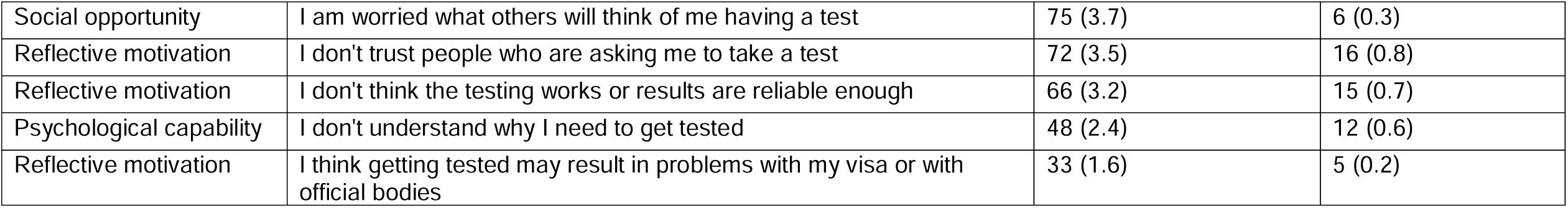
Study 2 prevalence of barriers in nationally representative sample (November 2020, n=2034)

| COM-B category | Barrier selected from x | Number of selections (%) | Number of people to rank it first (%) |
| --- | --- | --- | --- |
| Reflective motivation | I know what symptoms I have and don't believe they are COVID-19 ones e.g. hayfever/normal cold | 562 (27.6) | 200 (9.8) |
| Psychological capability | I'm not sure my symptoms are bad enough | 387 (19.0) | 109 (5.4) |
| Reflective motivation | It is unlikely I have COVID-19 because there aren't many cases in my area | 366 (18.0) | 104 (5.1) |
| Reflective motivation | I would prefer to self-isolate instead | 341 (16.8) | 73 (3.6) |
| Psychological capability | I'm not sure this symptom is one that needs testing | 306 (15.0) | 66 (3.2) |
| Reflective motivation | I think the test is painful | 276 (13.6) | 67 (3.3) |
| Reflective motivation | I'm worried I will catch COVID-19 myself whilst getting tested or on the way to the testing centre | 219 (10.8) | 34 (1.7) |
| Reflective motivation | I am worried about spreading my illness on the way to the testing centre | 207 (10.2) | 39 (1.9) |
| Reflective motivation | I will only get tested if my GP tells me I should | 187 (9.2) | 63 (3.1) |
| Reflective motivation | I have already got tested for these symptoms and was negative | 150 (7.4) | 34 (1.7) |
| Reflective motivation | I think the process of testing is too much effort | 148 (7.3) | 15 (0.7) |
| Reflective motivation | I don't want to take public transport | 143 (7.0) | 31 (1.5) |
| Psychological capability | I'm not sure how to get tested | 123 (6.0) | 27 (1.3) |
| Reflective motivation | I don't want to self-isolate after the test | 118 (5.8) | 16 (0.8) |
| Social opportunity | I am worried what others will think of me being positive for COVID-19 | 117 (5.8) | 29 (1.4) |
| Psychological capability | I have been told not to get tested even if I have symptoms | 113 (5.6) | 33 (1.6) |
| Reflective motivation | I had a bad experience when I got tested before | 89 (4.4) | 10 (0.5) |
| Reflective motivation | I have the symptoms but will wait for them to get worse first | 88 (4.3) | 9 (0.4) |
| Reflective motivation | I don't want to take up resources for testing so that others can't get tested | 88 (4.3) | 8 (0.4) |
| Reflective motivation | I don't want to hear that I'm positive for COVID-19 | 85 (4.2) | 5 (0.2) |
| Social opportunity | I am worried what others will think of me having a test | 75 (3.7) | 6 (0.3) |
| Reflective motivation | I don't trust people who are asking me to take a test | 72 (3.5) | 16 (0.8) |
| Reflective motivation | I don't think the testing works or results are reliable enough | 66 (3.2) | 15 (0.7) |
| Psychological capability | I don't understand why I need to get tested | 48 (2.4) | 12 (0.6) |
| Reflective motivation | I think getting tested may result in problems with my visa or with official bodies | 33 (1.6) | 5 (0.2) |

When we compared participants with low versus high health literacy, we found very similar issues arose for the top 10 barriers, covering reflective motivation and psychological capability. However, there were significant differences in the proportion of selections for the two groups. People with low health literacy were more likely to select certain capability issues: I’m not sure how to get tested, I’m not sure if this symptom needs testing; and motivation issues: I would prefer to self-isolate instead, I think the test is painful, I’m worried about spreading my illness, I’m worried I will catch COVID-19, I don’t want to hear that I’m positive, I don’t trust people who are asking me to take a test (Table 5).

**Table 5:**
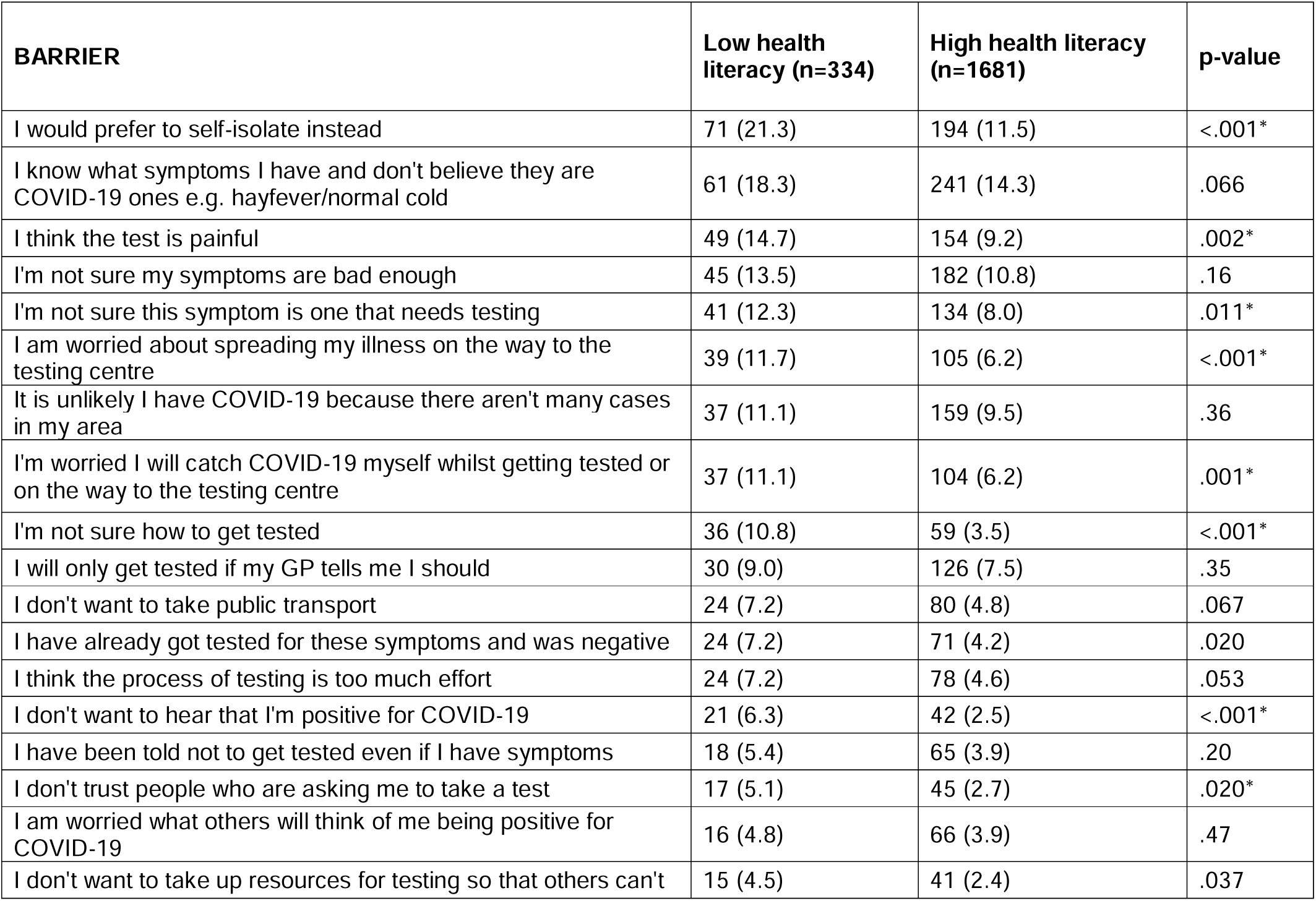
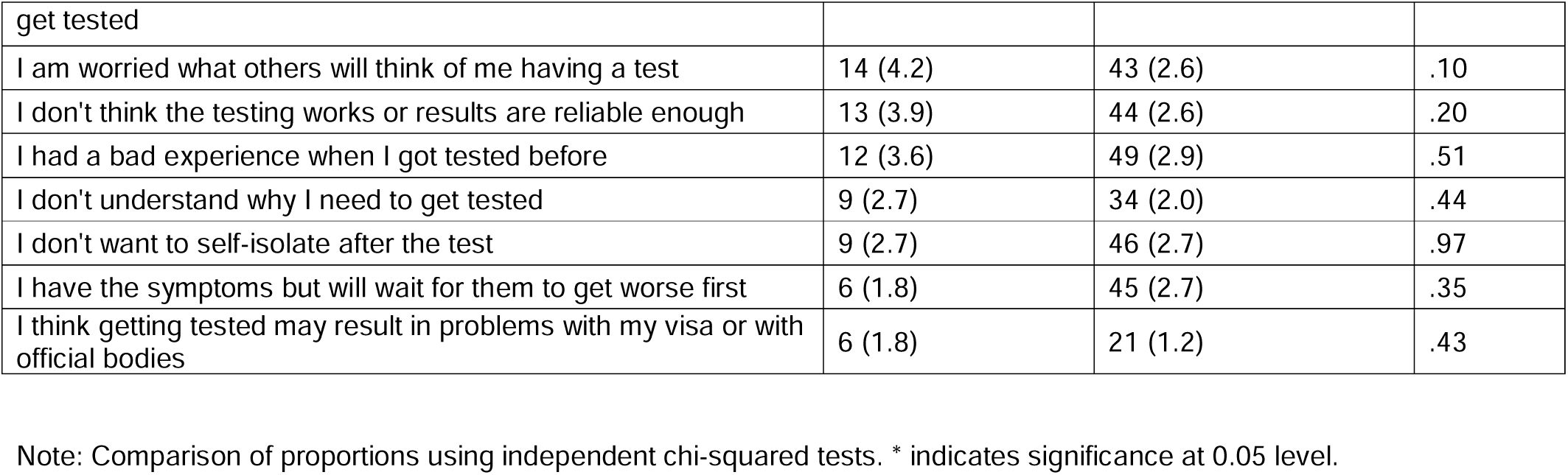
Study 2 frequency of barrier selection by health literacy level (November 2020, n=2034)

## DISCUSSION

These results provide new insights into behavioural barriers to COVID-19 testing, which is central to understanding and controlling the pandemic. Study 1 identified a wide range of barriers to COVID-19 testing that have not previously been described in the COVID-19 literature. These covered all three behavioural drivers in the COM-B model (capability, opportunity and motivation). Study 2 found that the motivation and capability barriers were far more prevalent than opportunity barriers in Australia. The top six most frequently selected barriers were the same as the most important barriers, covering psychological capability and reflective motivation. These issues were around misunderstanding COVID-19 symptoms, preferring to self-isolate rather than get a painful test, and believing there are few local cases. Many of these issues were more prevalent amongst people with lower health literacy. The large range of barriers identified in this study indicate that testing is a complex issue, so it requires a multifaceted approach to improve uptake depending on key barriers in different communities.

The most prevalent category of barriers in this study was motivation (e.g. don’t believe symptoms are COVID-19), which is likely to influence both testing intentions and behaviours. To address low intentions to test we need to identify and address the specific and wide-ranging concerns that individuals have. For some people, online symptom checkers could be a channel to provide tailored information, where users could be prompted for key concerns to link them to the most relevant information^21,22^, as opposed to searching for this in long ‘Frequently Asked Questions’ pages on government health websites. However, symptom checkers are not always reliable^23^, and may not be accessed by some groups such as culturally and linguistically diverse communities. Public health messaging could be better targeted to key motivation issues in different communities to address this. Our findings suggest a need to make it clear that self isolation means 14 days not just a few days while you have symptoms, so this is not seen as a good reason to avoid a painful test. We could also appeal to community benefits of testing, as this may be more motivating than individual risk in areas with low rates of COVID-19^24^. To address gaps between testing intentions and actual behaviour, people could be supported to plan in advance for the inconvenience of testing and self isolation; for example, managers could have a plan worked out in advance with their employees for how they will notify work and change shifts or work from home. Setting goals in advance has been shown to change various health-related behaviours^25,26^, and this strategy has been applied in Australia to help communities plan for bushfire management^27^.

For capability barriers (e.g. not sure which symptoms need testing), we know that government information about COVID-19 in Australia and in many countries is difficult to understand for the average person based on readability analyses^28,29^, and that communities with different language needs are not being adequately addressed^30^. We have also identified that certain groups are more susceptible to misinformation, such as younger people and men^19^. Addressing different information needs in the community may require using less traditional communication channels^31^ and different spokespeople to reach these groups, such as social media influencers^32^.

Although the Australian testing system is free and widely accessible, there are still important opportunity barriers particularly around financial losses and work expectations that require system changes. The state of Victoria addressed this issue during an outbreak in 2020 by providing financial support for those who were self isolating following a COVID-19 test, to support casual workers and others without access to sick pay^33^. There are also social opportunity issues where stigma and perceived judgment by others could prevent individuals from getting tested, which we have seen in other infectious disease conditions such as HIV^34^.

The study occurred in a context where risk of COVID-19 has been relatively low compared to many other countries worldwide. Testing behaviour has been a key part of identifying and controlling emerging COVID-19 outbreaks. This will likely become more relevant in countries where vaccine rollout enables the current large number of new cases to be contained. Our findings from June 2020 are very similar to September results from national flu tracking data, which show 51% of people who experienced COVID-19 symptoms did not get tested^35^. We found 51% of people did not “strongly agree” they would get tested, but most people agreed to some extent and addressing perceived barriers may improve this overall positive orientation to COVID-19 testing. Another survey conducted in Australia in August 2020 found much higher rates of testing avoidance, and reported 85% of respondents who had symptoms had not been tested^36^. The effect of vaccine rollout on testing intentions in Australia remains to be seen, with only a small proportion of the population currently vaccinated and none at the time of data collection^37^.

### Future research

We have registered a trial to test a tailored intervention to address the capability and motivation issues identified in this study^38^ (ACTRN12620001355965), but broader system approaches are also needed to address opportunity issues. This may include working with the media to avoid the identification and stigmatisation of individuals with positive COVID-19 test results, which has repeatedly happened where a single case is linked to a new outbreak^39^. Our findings provide a starting point for other surveys to identify and address local testing barriers. Media reports and anecdotal data from frontline health professionals may be a useful way to quickly identify emerging local issues that could supplement our list of testing barriers to make it more relevant to local communities.

### Strengths and limitations

Our initial study only provided a limited list of testing barriers proposed by the NSW Department of Health, but this sample was not representative and did not cover a large number of “other” responses. We addressed this in our second study with a nationally representative sample and a full list of barriers. However, even the second study was not representative of all community groups, particularly those from culturally and linguistically diverse backgrounds which has been identified as a key area of need in Australia. We are currently conducting a separate survey with these groups using interpreters to conduct the survey in preferred languages, as a partnership with Western Sydney Local health District. The findings may not be generalisable to other countries, particularly where COVID-19 prevalence is too high for a test and trace approach to be feasible, and where opportunity issues such as cost or physical access to testing is a problem. However, understanding COVID-19 testing barriers can help all countries better prepare for the next pandemic where a test-trace-isolate system can be used.

## Conclusion

This paper uncovers a wide range of barriers to COVID-19 testing that had not previously been identified in the COVID-19 prevention literature. The prevalence estimates show that motivation and capability issues need to be addressed in Australia in order to increase testing behaviour, particularly for groups with lower health literacy. Interventions to improve testing uptake need to identify key barriers for target populations, and specifically address these using evidence-based behaviour change techniques. Our findings support broader advice from international experts in behaviour change, highlighting the importance of diagnosing behavioural barriers in order to increase adherence with COVID-19 prevention behaviours^2^.

## Data Availability

Please for information about the data

## ACKNOWLEDGEMENTS

We thank the participants of the longitudinal COVID-19 survey for their ongoing participation in this research. This study was not specifically funded, but in-kind support was provided by authors with research fellowships.

CB is supported by a National Health and Medical Research Council (NHMRC)/Heart Foundation Early Career Fellowship (#1122788).

RD is supported by a University of Sydney fellowship (#197589).

KM is supported by a National Health and Medical Research Council (NHMRC) Principal Research Fellowship (#1121110).

